# Variation in *ERAP2* has opposing effects on severe respiratory infection and autoimmune disease

**DOI:** 10.1101/2022.11.04.22281942

**Authors:** Fergus Hamilton, Alexander Mentzer, Tom Parks, J Kenneth Baillie, George Davey Smith, Peter Ghazal, Nicholas J Timpson

## Abstract

*ERAP2* is an aminopeptidase involved in immunological antigen presentation. Genotype data in human samples from before and after the Black Death, an epidemic due to *Yersinia pestis*, have marked changes in population allele frequency of the common single nucleotide polymorphism (SNP) rs2549794. This SNP in strong linkage disequilibrium with a key splicing SNP in *ERAP2* (rs2248374) and this suggests that variation at *ERAP2* may be relevant for protection from infection. rs2549794 is also associated with Crohn’s disease and findings imply balancing selection between infection and autoimmune disease at this locus. There have been no large-scale prospective case-control studies of variation at *ERAP2* and infection.

**Methods:** This study aimed to explore the association between variation at *ERAP2* and a) infection, b) autoimmune disease, and c) parental longevity as a proxy for lifespan. Genome Wide Association Studies (GWAS) of these outcomes were identified in contemporary cohorts (UK Biobank, FinnGen, and GenOMICC). Effect estimates were extracted for rs2549794 and rs2248374. Additionally, *cis* expression and protein quantitative trait loci (QTLs) for *ERAP2* were used in Mendelian randomisation analyses.

**Results:** Across all cohorts, the T allele (minor allele frequency of 0.4-0.5) of rs2549794 showed evidence of association with respiratory infection (odds ratio; OR for pneumonia 1.03; 95% CI 1.01-1.05; p = 0.014). Effect estimates were larger in bacterial rather than viral infection and larger for more severe phenotypes (OR for critical care admission with pneumonia 1.08; 95% CI 1.02-1.14, p = 0.008, OR for death from pneumonia 1.07; 95% CI 1.01-1.12; p = 0.014). In contrast, opposing effects were identified for Crohn’s disease (OR 0.86; 95% CI 0.82-0.90, p = 8.6 × 10^−9^) and type 1 diabetes (OR 0.95; 95% CI 0.90-0.99, p = 0.02). No strong evidence for association was identified for sepsis. Carriage of the T allele was associated with increased age of parental death (beta in Z-scored years across both parents age at death 0.01, 95% CI 0.004-0.017, p = 0.002). Similar results were identified for rs2248374.

In Mendelian randomisation analyses, increasing transcription or protein levels of *ERAP2* were strongly associated with protection from respiratory infection, with opposing effects identified on Crohn’s disease and type 1 diabetes. Increased expression of *ERAP2* was associated with reduced parental longevity.

**Conclusions:** Variation at *ERAP2* is associated with severe respiratory infection in modern societies, with an opposing association with Crohn’s disease and type 1 diabetes. These data support the hypothesis that changes in allele frequencies in *ERAP2* observed at the time of the Black Death reflect protection from infection, and suggest ongoing balancing selection at this locus driven by autoimmune and infectious disease

## Introduction

*ERAP1* and *ERAP2* both code for aminopeptidases that are critical in presentation of antigen by professional antigen presenting cells and are linked with the Human Leukocyte Antigen (HLA) response to infection.^1–4^ Variation at these genes has been associated with both infection and autoimmune disease, with the most compelling association reported between variation at *ERAP2* and inflammatory bowel disease (particularly Crohn’s disease), and *ERAP1* with ankylosing spondylitis.

Other studies have suggested a role for an association with *ERAP2* and infection, although studies have been small. Additionally, it is thought that these genes are under balancing selection, with ERAP2 having two, almost equally common haplotypes worldwide (Haplotype A and Haplotype B).^5–7^ Haplotype A encodes the full ERAP2 protein, whereas Haplotype B has a premature stop codon leading to nonsense mediated decay, and reduced levels of ERAP2 protein.^8^

A recent analysis by Klunk et al of human genomes from before, during, and after the Black Death in Europe, an epidemic caused by *Yersinia pestis*, identified a large change in allele frequency over this period in a common SNP (rs2549794-T) in *ERAP2*.^8^ This SNP is in linkage disequilibrium with the known splicing variant (rs2248374-G) that leads to transcription of the specific haplotype (Haplotype B), that is associated with production of a truncated ERAP2 protein which undergoes nonsense mediated decay and appears to affect antigen presentation.^5^ The new work has suggested that rs2549794 is associated with the immune response, with carriers of the putative protective (C) allele having a five-fold higher *ERAP2* expression in both unstimulated and *Y*.*pestis* challenged macrophages, altered gene expression patterns in multiple other immune cells, and increased production of the full length ERAP2 protein in macrophages. Additionally, the rs2549794-C allele has been associated with increased odds of Crohn’s disease in other studies.^9^ Therefore, the study concludes that variation at *ERAP2* (in particular, the C allele at rs2549794) is probably associated with protection from *Y*.*pestis* and suggest that the balancing selection at this locus is likely driven by a balance between autoimmune disease (or other negative consequences of this SNP) and protection from severe infection.

This conclusion remains a hypothesis and there is little data on whether variation at *ERAP2* (and at this specific SNP) associates with severe infection and other autoimmune diseases outside Crohn’s in contemporary datasets.^10,11^ In this study, we aim to test this hypothesis by identifying whether variation at *ERAP2* is associated with serious infection (sepsis and respiratory infection, autoimmune disease and age at parental death (as a proxy for potential effects on longevity) using both the SNP identified by Klunk et al^8^, and by using expression^12,13^ and protein^14^ quantitative trait loci for *ERAP2*.

## Results

We identified Genome-wide association study (GWAS) summary statistics for 7 respiratory infection outcomes, 5 sepsis outcomes, 4 autoimmune disease outcomes, and 3 (mother, father, both) parental longevity-related outcomes. **Table 1** lists details of each study, with descriptions of the cohorts in the methods.

**Table 1:**
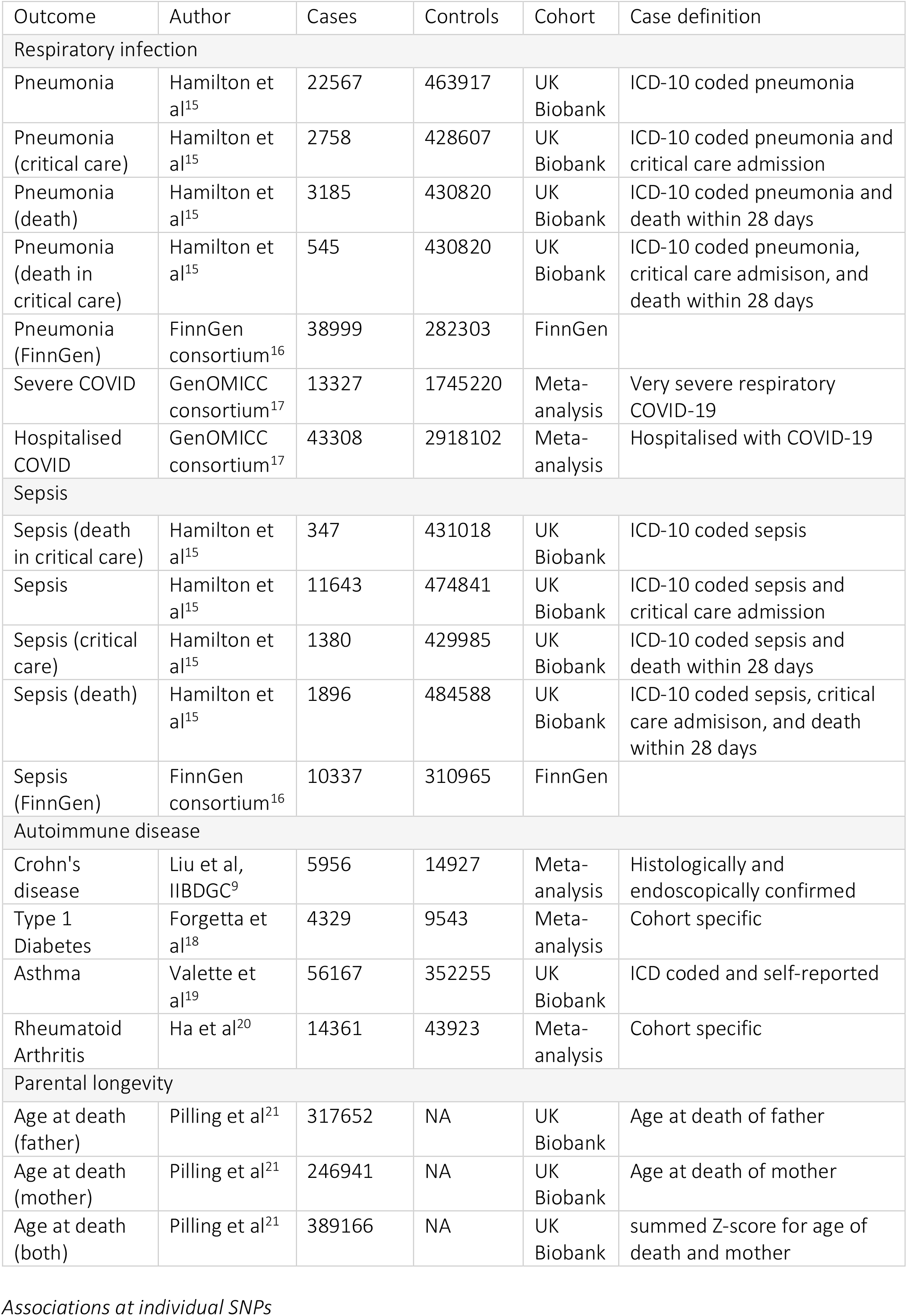
Studies included in this analysis

### Associations at individual SNPs

In our initial analysis in UK Biobank, we focussed on rs2549794. This SNP has a minor allele frequency of between 0.4-0.5 in modern European cohorts. The T allele of rs2549794 was found to be associated with increased odds of respiratory infection (**Figure 1**), but not sepsis. Effect sizes increased with severity of disease, with the largest effects for death from critical care pneumonia (OR 1.11; 95% CI 0.98 – 1.26, p = 0.094), and for critical care admission with pneumonia (OR 1.09; 95% CI 1.02 – 1.14, p = 0.008).

**Figure 1:**
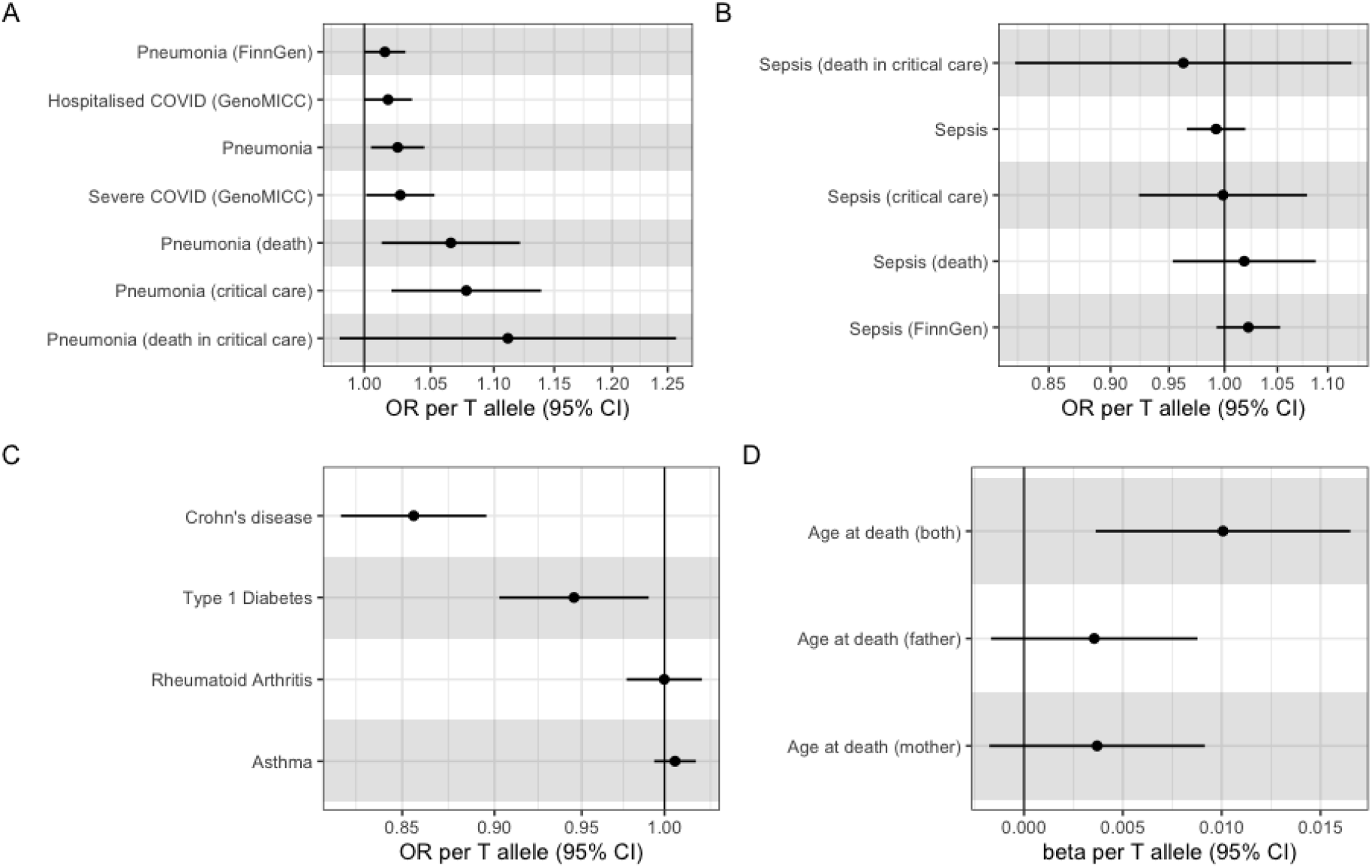
Associations between rs2549794 and outcomes in each GWAS dataset. Odds ratio for each outcome presented, with 95% confidence intervals, except for parental longevity, where the beta is presented with a 95% confidence interval. A) respiratory infection B) sepsis, C) Autoimmune disease D) parental longevity

The effect size on likely bacterial respiratory infection was larger than that on viral infection (COVID-19 in GenoMICC)^17^, although all effects were modest in size (OR < 1.15). As expected, this allele was associated with protection from Crohn’s disease (OR 0.86; 95% CI 0.82 – 0.90, p = 8.6 × 10^−9^) and we identified a protective association with type 1 diabetes (OR 0.95; 95% CI 0.90-0.99, p = 0.02, all studies in **Table 1**). In existing data, we were unable to resolve any association with asthma or rheumatoid arthritis. There was evidence of an association with parental longevity, with each additional T allele being associated with increased age at parental death (beta 0.01 on Z-scored combined age of death of parents, 95% CI 0.004-0.017, p = 0.002), with no difference between maternal and paternal estimates. **Table 2** show results of this analysis. Associations at rs2248374, the splicing variant in linkage disequilibrium with the above SNP, were very similar as would be expected given the LD in European populations (**Supplementary Figure 1, Supplementary Table 1**).

**Table 2:** Associations between rs2549794 T allele and outcomes in each GWAS dataset. Odds ratio for each outcome presented, with 95% confidence intervals, except for parental longevity, where the raw beta is presented with a 95% confidence interval. NR = not reported. EAF = Effect allele frequency

| Outcome | OR/beta | Lower 95% CI | Upper 95% CI | P value | EAF |
| --- | --- | --- | --- | --- | --- |
| **Respiratory Infection (OR reported)** |  |  |  |  |  |
| Pneumonia | 1.025 | 1.005 | 1.045 | 0.014 | 0.565 |
| Pneumonia (critical care) | 1.078 | 1.020 | 1.139 | 0.008 | 0.567 |
| Pneumonia (death) | 1.066 | 1.013 | 1.121 | 0.014 | 0.565 |
| Pneumonia (death in critical care) | 1.111 | 0.982 | 1.258 | 0.094 | 0.567 |
| Pneumonia (FinnGen) | 1.015 | 1.000 | 1.031 | 0.048 | 0.621 |
| Severe COVID (GenoMICC) | 1.027 | 1.001 | 1.053 | 0.038 | NR |
| Hospitalised COVID (GenoMICC) | 1.018 | 1.000 | 1.036 | 0.053 | NR |
| **Sepsis (OR reported)** |  |  |  |  |  |
| Sepsis (death in critical care) | 0.963 | 0.824 | 1.125 | 0.631 | 0.567 |
| Sepsis | 0.992 | 0.966 | 1.019 | 0.567 | 0.565 |
| Sepsis (critical care) | 0.999 | 0.924 | 1.079 | 0.970 | 0.567 |
| Sepsis (FinnGen) | 1.022 | 0.993 | 1.053 | 0.144 | 0.622 |
| Sepsis (death) | 1.018 | 0.953 | 1.088 | 0.590 | 0.565 |
| **Autoimmune disease (OR reported)** |  |  |  |  |  |
| Asthma | 1.001 | 0.999 | 1.002 | 0.253 | 0.562 |
| Type 1 Diabetes | 0.945 | 0.903 | 0.990 | 0.017 | 0.570 |
| Rheumatoid Arthritis | 0.990 | 0.968 | 1.012 | 0.380 | NA |
| Crohn's disease | 0.856 | 0.818 | 0.896 | 0.000 | 0.588 |
| **Age at death (beta reported)** |  |  |  |  |  |
| Age at death (father) | 0.004 | -0.002 | 0.009 | 0.170 | 0.567 |
| Age at death (mother) | 0.004 | -0.002 | 0.009 | 0.190 | 0.567 |
| Age at death (both) | 0.010 | 0.004 | 0.0017 | 0.002 | 0.567 |

### Mendelian randomisation

Previous work has suggested that important variation (such as at rs2248374) at *ERAP2* acts via alternative splicing leading to altered RNA and protein levels.^1,5,8,22^ We therefore went on to perform Mendelian randomisation using expression and protein quantitative trait loci (QTL) *cis* for *ERAP2*, to identify if altered levels of ERAP2 transcripts or protein altered the odds of each outcome. We identified independent (r^2^ > 0.1, in the European population of 1000 Genomes Project^23^) SNPs that were associated (p < 5 × 10^−8^)with either a) whole blood RNA expression, b) lung tissue RNA expression, or plasma protein levels of ERAP2. Details on derivation of these are in the methods, with a list of all included SNPS in **Supplementary Table S2.**

In inverse variance weighted meta-analysis, increased whole blood *ERAP2* expression was associated with protection from respiratory infection (**Figure 2**). Again, this effect was most pronounced for the most severe infection, with the largest effect estimates for patients who died or were critically unwell with pneumonia: OR for death from pneumonia 0.94; 95% CI 0.92 – 0.96, p = 4 × 10^−7^; OR for pneumonia 0.98; 95% CI 0.98-0.99, p = 7 × 10^−4^). Effect sizes were much larger in pneumonia (largely bacterial) than in COVID-19. As in the individual SNP analyses no effect was identified on sepsis related outcomes, and opposing effects were identified on type 1 diabetes (OR 1.05; 95% CI 1.03-1.08, p = 3 × 10^−5^) and Crohn’s disease (OR 1.09, 95% CI 1.07-1.11; p = 2.7 × 10^−16^). Increased *ERAP2* expression was associated with increase age at death in both men and women, with a combined estimate of 0.007; 95% CI 0.003 - 0.010; p = 3.8 × 10^−5^).

**Figure 2:**
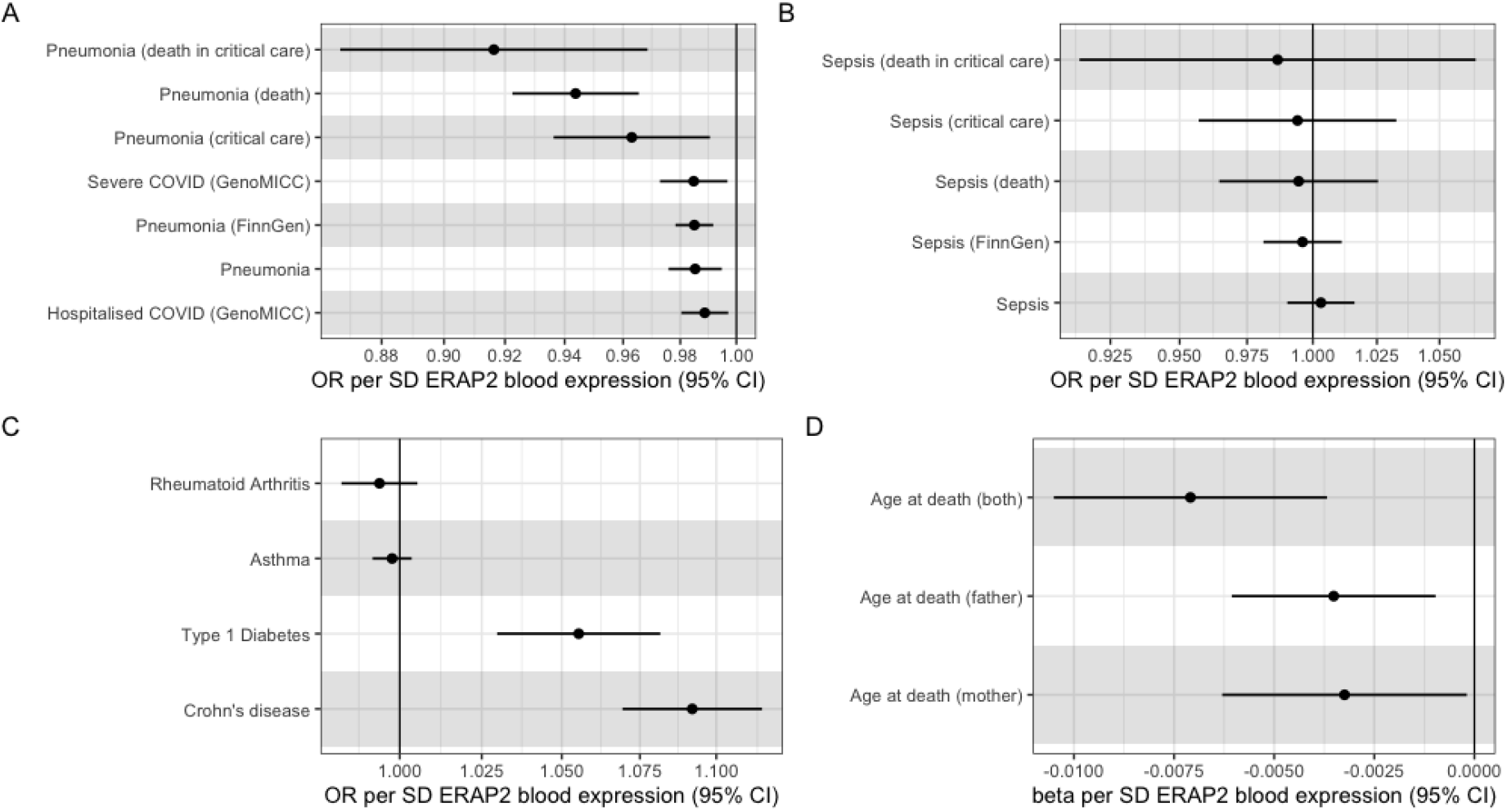
Inverse variance weighted MR estimates for the effect of *ERAP2* whole blood expression on outcomes. Effect estimates are on the scale of normalised gene expression. Results for A) respiratory infection B) sepsis, C) Autoimmune disease D) parental longevity

Using lung tissue specific eQTLs, effect sizes were similar, although the strength of the association was weaker, reflecting in part the fewer number of SNPs in these analyses (**Supplementary Figure 2**). Analyses using protein quantitative trait loci were again similar (**Supplementary Figure 3**), although associations with COVID were null, and other associations are weaker. All QTL results are reported in **Supplementary Tables 3-5**, while **Supplementary Data 1** reports weighted median and MR Egger results for each analysis. These results were similar, but more imprecise than our primary analysis. As a final sensitivity analysis, we repeated all analyses using a more stringent independence cut off (r^2^ < 0.01). These results were similar, with inverse variance weighted MR analyses for whole blood *ERAP2* expression reported in **Supplementary Figure 4**, and full results in **Supplementary Data 1.**

## Discussion

In this study, we show that variation at *ERAP*2 is associated with susceptibility to respiratory infection in the present day. In line with a recent study using ancient DNA^8^, we show evidence suggesting that the C allele of rs2549794 is protective against respiratory infection, with evidence of increasing protection against severe disease. Although the effect size of the T allele on respiratory infection is modest (OR 1.05-1.1), this plausibly represents a large attenuation of the “wild type” effect of this locus, given modern treatments for respiratory infection (e.g. antimicrobials) and other public health measures that are likely to blunt the genetic effects. In contrast, we identified an opposing effect on Crohn’s disease and type 1 diabetes, providing some support for the hypothesis of balancing selection at this locus.^5–8^ Using parental death data from UK Biobank, we identified evidence of reduced parental lifespan with carriage of the C allele at rs2549794.

*ERAP2* is characterised by two Haplotypes (A + B), with Haplotype B leading to reduced expression of ERAP2 and low amounts of truncated ERAP2 protein.^5,8^ This reduced amount and function of ERAP2 alters the diversity of antigens presented by HLA, and is plausibly likely to reduce the quality of immune response to certain pathogens.^4,24^ Haplotype B is characterised by the presence of rs2248374-G. Therefore, we would expect that reduced expression and translation of *ERAP2* would have similar effects to rs2248374-G.

Given that, our Mendelian randomisation analyses using expression and protein quantitative trait loci at *ERAP2* are highly concordant with the individual SNP data and underlying biology, showing evidence of reduced odds of severe respiratory infection with increasing levels of *ERAP2* expression or protein. The same inverse association with Crohn’s disease and type 1 diabetes was identified. The evidence of association was strongest with whole blood eQTL data, although we saw effects using all three of our instruments (whole blood and lung eQTL, plasma pQTL).

We saw no strong evidence for association with data based on sepsis related clinical coding. This is interesting, as most data point to the bubonic form of *Y*.*pestis* as the predominant feature of the Black Death, although the speed of travel of the epidemic has made some consider whether pneumonic plague was a more plausible candidate..^8^ It is also important to recognise that sepsis represents a dysregulated response to infection^25^, and is highly heterogenous over even the last 50 years, perhaps explaining the lack of association.^26^ We did, however, see an association with reduced parental longevity with the C allele and with increased *ERAP2* expression. It is possible to speculate about the cause of this, and whether this is entirely represented by an increased rate of Crohn’s disease and Type 1 diabetes. As both Crohn’s disease and Type 1 diabetes are known to reduce fertility, it may be that some balancing selection is driven by this reduced fertility, although this requires further investigation.^27–29^

Although it has been long suspected that *ERAP2* has a role in protection from infection (reviewed recently^2^), with known roles in both antigen presentation and shaping the cytokine response^30^ there are no studies as far as we are aware there are that have robustly identified genetic associations on the population level. There are, however, some candidate gene studies reported, although given the small sample sizes their relevance to our analyses is unclear.^1,2,6,11,31^

The major limitation of this work – in line with other genetic association studies in infection – is the challenge in defining cases of infection, which differ between and across studies. Additionally, there are assumptions and challenges of interpreting MR which are discussed extensively elsewhere,^22,32^ but are more challenging at a locus such as *ERAP2* which has complex biology and a complex genetic background.^5^

There are three clear implications of the. Firstly, this data confirms the hypotheses raised by other studies:^5–8^ there is likely a balancing effect at *ERAP2* driven by protection from infection with increased risk of Crohn’s disease and type 1 diabetes. Secondly, therapeutics to target *ERAP1* and *ERAP2* are in development (to target Crohn’s disease and cancer^33^). These data suggest targeting *ERAP2* may lead to increased rates of respiratory infection; and trials should measure and quantify infection outcomes. Finally, these findings show the potential opportunities in contemporary infection genetics: identifying and prioritising potential gene targets using DNA from samples that are centuries old, quantifying associations in modern datasets, and identifying potential off-target effects.

## Conclusion

Variation in *ERAP2* is associated with respiratory infection. In particular, the C allele of rs2549794 appears to be protective, with increasing effect estimates with increasing severity of infection. This is consistent with the protective effect identified in analyses of genomes around the time of the Black Death. In supporting Mendelian randomisation analyses, increased expression of *ERAP2* is protective for respiratory infection, but increase the odds of some autoimmune diseases and are associated with reduced parental longevity. These results suggest ongoing balancing selection at this locus driven by autoimmune and infectious disease.

## Materials and methods

### Study design

For this study, we aimed to assess whether variation at *ERAP2* was associated with severe infection, autoimmune disease, and parental longevity, as a proxy for an effect on lifespan.

### Infection outcomes

For our pneumonia and sepsis outcomes we utilised our previously performed case-control GWAS of sepsis and pneumonia from UK Biobank.^15^ UK Biobank is a large (n ∼ 500,000) cohort of older (>50) UK residents who were recruited between 2002 and 2009, and who have linked electronic health record and mortality data. Details of the cohort are available elsewhere.^34^

For this study we included data on both incidence outcomes (presence of disease), mortality outcomes (death within 28 days of disease), and critical care related outcomes (critical care admission with the disease).

Cases were identified by the presence of ICD-10 coding in linked hospital data, and deaths from nationally linked mortality data.^34^ Comparisons were performed against all other participants in UK Biobank (e.g. death within 28 days of pneumonia vs all other participants in UK Biobank). All analyses were performed in participants of European ancestry (N ∼ 350,000). Further details on case definition, analysis approach, and GWAS methodology are with the original publication^15^ and elsewhere^35^, while GWAS are available at the MRC-IEU GWAS database.^36^

Additionally, we identified matching GWAS for disease incidence from the FinnGen Round 7.^16^ FinnGen is a similar, large, population cohort in Finland, with linked genetic and health outcome data. Case-control GWAS have been performed for ICD-10 coded hospital admission, with details available at the FinnGen website.^16^

Finally, we identified COVID-19 outcomes from the GenoMICC^17^ (Round 3) consortium. GenoMICC is a as an open-source international study recruiting critically ill patients starting in 2015, with this analysis focussing on the comparison between hospitalised patients with COVID-19 and the general population (Hospitalised COVID-19), and comparisons between severely unwell patients with COVID-19 and the general population (Severe COVID-19).

### Autoimmune disease outcomes

We focussed on four common autoimmune diseases which have existing GWAS summary statistics available: Type 1 diabetes, asthma, Crohn’s disease, and rheumatoid arthritis. For type 1 diabetes, we utilised summary statistics from a recent meta-analysis of 12 European cohorts.^18^ For asthma, we utilised a recent GWAS in UK Biobank using a broad definition of Asthma.^19^ For Crohn’s disease we utilised a GWAS meta-analysis performed in a European population from the International Inflammatory Bowel Disease Genetics Consortium.^9^ Finally, for rheumatoid arthritis, we utilised summary statistics from a recent meta-analysis of rheumatoid arthritis.^20^

### Parental longevity outcomes

Parental longevity was chosen as a proxy measurement to identify potential effects of exposures on lifespan. For this analysis, we utilised GWAS summary data from a recent GWAS by Pilling et al in UK Biobank.^21^ On recruitment to UK Biobank, participants were asked what age both parents had died at, and this data was used to generate age at death GWAS. Exclusion criteria were applied to exclude extremely early death, with details in the manuscript. To generate the combined across parents age at death data, each age was converted into a Z-score, and then this was summed to generate a combined phenotype.

### Analytic approach

We had two analytic approaches. Firstly, we focussed on the SNP identified by Klunk et al^8^ (rs2549794), and the linked splicing variant (rs2248374), and performed association analyses for the presence of these SNP in each outcome dataset.

Secondly, we performed Mendelian randomisation^22^ using expression and protein quantitative trait loci (QTLs) *cis* to *ERAP2*. Mendelian randomisation uses SNPs as instrumental variables to explore the association between an exposure (e.g. *ERAP2* expression) and an outcome (e.g. pneumonia).^22^ Mendelian randomisation is a form of instrumental variable analysis, that under certain assumptions can provide causal estimates of the effect of an exposure on an outcome. These assumptions are that the genetic instruments are associated with the risk factor of interest, were independent of potential confounders, and could only affect the outcome through the risk factor and not through alternative pathways (that is, through pleiotropy).^22^ In this study, our instruments were all *cis* acting QTLs. We identified expression QTLs from whole blood (from eQTLGen^37^), and eQTLs from lung tissue from the GTEx consortium^12^. Protein QTLs were identified from GWAS performed by the DECODE consortium.

For all of these exposures, SNPs robustly (p < 5x 10^−8^) associated with the exposure (expression or protein levels) within 300kb of *ERAP2* were extracted. SNPs were pruned to ensure independence (r^2^ = 0.1). Effect alleles were harmonised with the outcome datasets. MR was then performed for each SNP individually and results meta-analysed via inverse variance weighting. Other meta-analytical methods are reported as sensitivity analyses. As a final sensitivity analyses, we pruned the SNP’s further using a r^2^ threshold of 0.01, and re-ran analyses.

### Software

Analysis was performed using R v 4.0.4 (R Foundation for Statistical Computing, Vienna). Data wrangling was performed using the tidyverse, and plotting using ggplot2.^38^ Two Sample Mendelian randomisation was performed using the TwoSampleMR package in R.^36^

## Supporting information

S1 STROBE-MR

## Data Availability

All data available to replicate this analysis is publicly available. For ease, curated data is available at the authors GitHub (https://github.com/gushamilton/ERAP2) allowing ease of replication.

## Guidelines

This study is reported in line with the STROBE-MR guidance.^39^ (**Supplement S1**)

