## Supplementary Figures for "Variation in *ERAP2* has opposing effects on severe respiratory infection and autoimmune disease"

**Supplementary Figure 1:** Association between rs2248374-G and outcomes in each GWAS dataset. A) respiratory infection B) sepsis, C) Autoimmune disease D) parental longevity

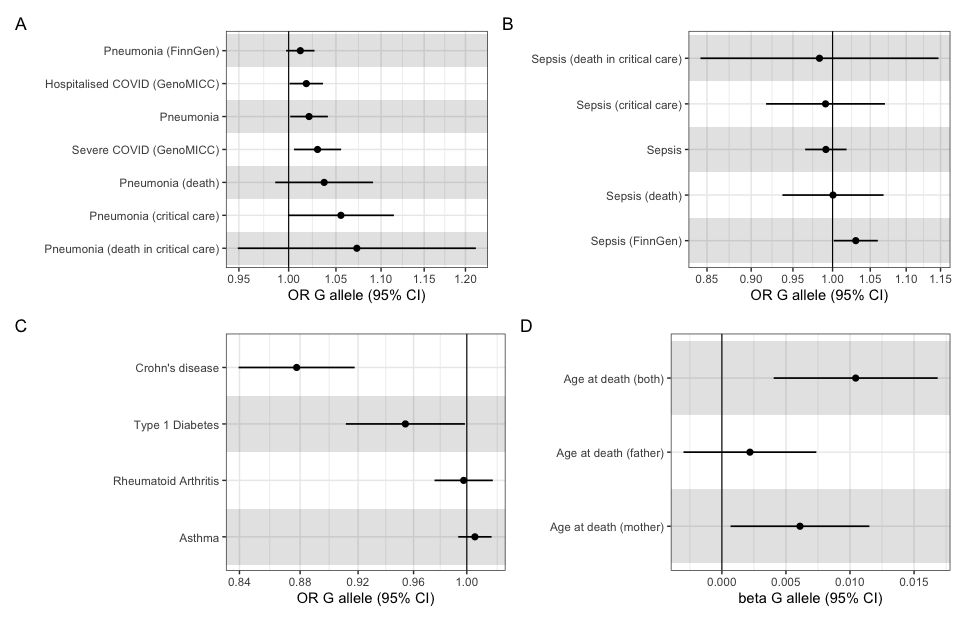

**Supplementary Figure 2:** Association between *ERAP2* lung tissue expression and outcomes in each GWAS dataset. Estimates generated from IVW MR. A) respiratory infection B) sepsis, C) Autoimmune disease D) parental longevity

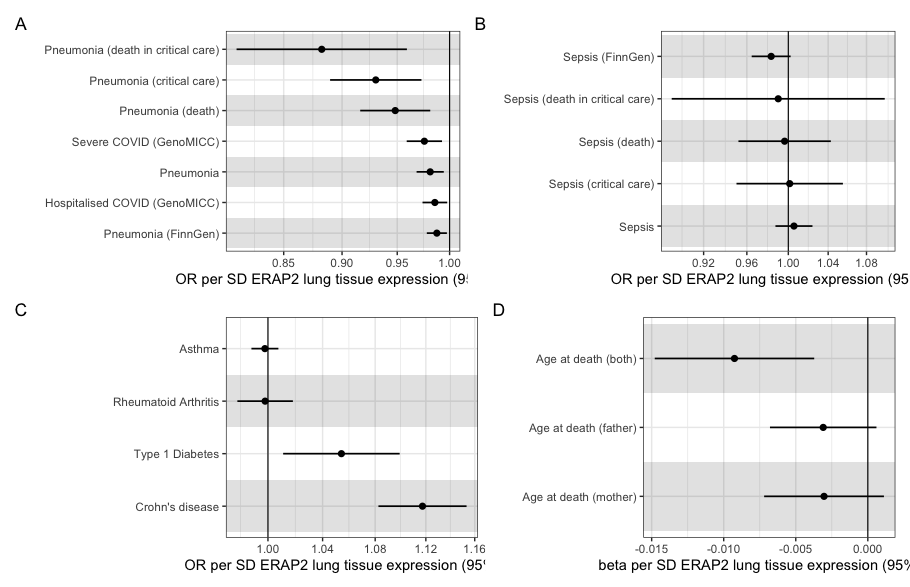

**Supplementary Figure 3:** Association between *ERAP2* protein levels and outcomes in each GWAS dataset. Estimates generated from IVW MR. A) respiratory infection B) sepsis, C) Autoimmune disease D) parental longevity

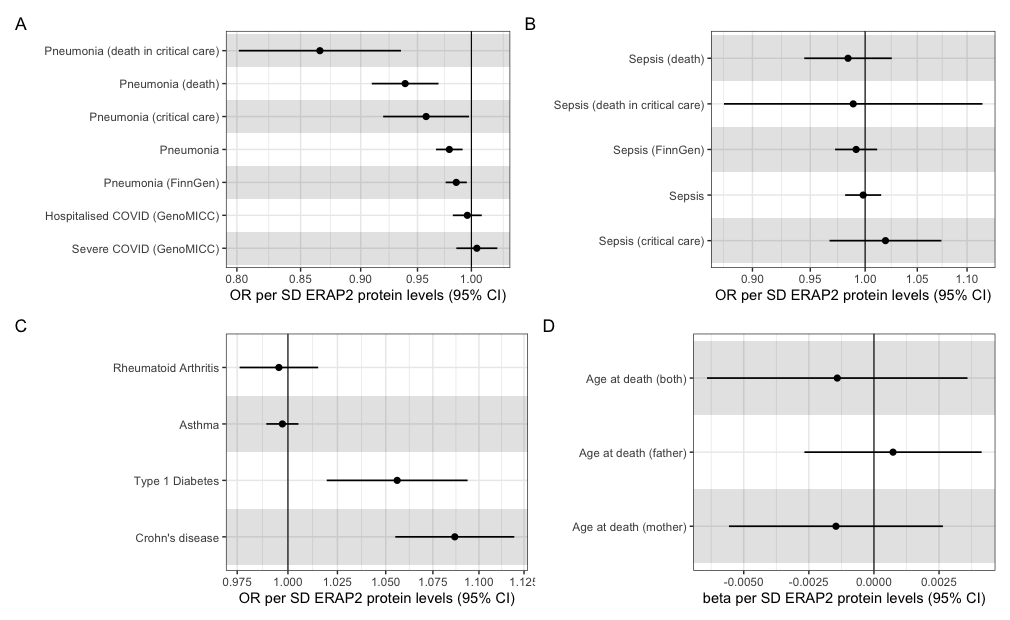

**Supplementary Figure 4:** Association between *ERAP2* whole blood gene expression with SNP’s clumped at a threshold of r2 <0.01) and outcomes in each GWAS dataset. Estimates generated from IVW MR. A) respiratory infection B) sepsis, C) Autoimmune disease D) parental longevity

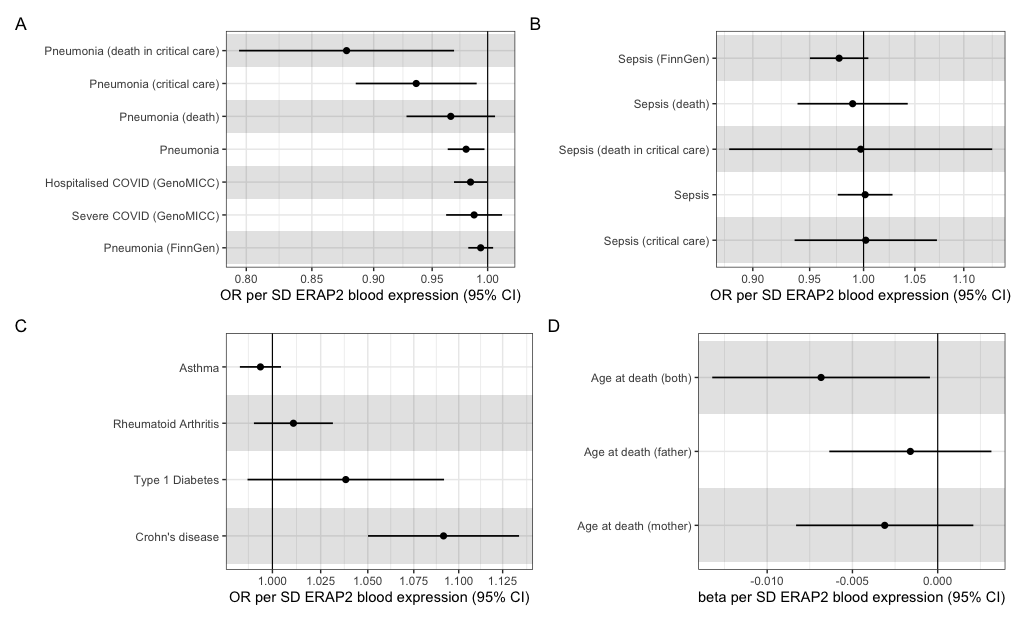
